# Longitudinal Study on Changes in COVID-19 Vaccination Intentions in Benin and Senegal: Insights from Generalized Estimating Equations (GEE)

**DOI:** 10.1101/2025.04.16.25325940

**Authors:** Ibrahima Gaye, Adama Faye, Valery Ridde, Avahoundje E. Martin

## Abstract

**Introduction:** The COVID-19 pandemic led to strict measures and the rapid deployment of vaccines, which were adopted in Benin and Senegal. This longitudinal study employs Generalized Estimating Equations (GEE) to analyze the evolution of vaccination intent and its determinants. By examining attitudes, risk perceptions, and social influence, it provides insights into adherence dynamics and helps guide vaccination strategies in West Africa.

**Methods:** This is a longitudinal, descriptive, and analytical study. The sample includes 546 Beninese, and 319 Senegalese individuals aged 18 and above, selected using marginal quotas. Data collection was conducted through Random Digit Dialing (RDD) using a questionnaire based on the Theory of Planned Behavior (TPB) and the Health Belief Model (HBM). The influence of factors was assessed using a GEE model.

**Results:** The evolution of vaccination intent between the two data collection phases was more pronounced in Senegal (+12.5 points, p=0.000) than in Benin (+5.0 points, p=0.089). The increase was significant among women (+19.9 points, p=0.000) and individuals under 25 years old (+15.6 points, p=0.030), whereas in Benin, this younger group showed a decline (–11.5 points, p=0.009), with only those aged 25-59 years demonstrating an increase (+11.7 points, p=0.011). In both countries, individuals surveyed during the second phase had a significantly higher likelihood of expressing vaccination intent (Benin: OR = 6.9, p < 0.001; Senegal: OR = 5.0, p < 0.001).

Additionally, the increase in vaccination intent was influenced by common factors such as perceived benefits (Benin: OR = 1.191; Senegal: OR = 1.412), social influence (Benin: OR = 1.377; Senegal: OR = 1.310), and favorable attitudes toward vaccination (Benin: OR = 1.266; Senegal: OR = 1.254). However, differences emerged: in Benin, perceived efficacy (OR = 1.287) and behavioral control (OR = 1.203) were significant, whereas in Senegal, concerns about vaccine safety posed a greater barrier (OR = 0.685 compared to 0.781 in Benin).

**Conclusion:** This study highlights both common and specific determinants of vaccination intent in Benin and Senegal, particularly social influence and perceived vaccine benefits. However, contextual differences exist, such as the greater importance of perceived efficacy in Benin and the more pronounced impact of safety concerns in Senegal. These findings underscore the need to tailor communication strategies and strengthen public trust to improve vaccine uptake.

## INTRODUCTION

On March 11, 2020, the World Health Organization (WHO) declared COVID-19 a pandemic and urged countries to take immediate measures to limit the spread of the infection while adhering to the International Health Regulations (2005) [1]. Government responses included strict measures such as bans on public gatherings, social distancing, and lockdowns. Simultaneously, the rapid development of COVID-19 vaccines emerged as a crucial solution to contain the virus and achieve herd immunity [2, 3].

The WHO quickly granted emergency use authorization for several vaccines, including Pfizer (December 31, 2020), AstraZeneca (February 15, 2021), Johnson & Johnson (March 12, 2021), and Sinopharm (May 7, 2021) [4]. In this context, the governments of Benin and Senegal launched their vaccination campaigns on February 23 and March 29, 2021, respectively.

In a previous article [5], we explored the determinants of vaccination intent through a cross-sectional study conducted in Benin and Senegal using structural equation modeling (SEM). This analysis examined the influence of several factors, including individual attitudes toward vaccination, risk and safety perceptions, and the impact of social influence. The findings highlighted contextual variations between the two countries, suggesting that these different dimensions interact differently depending on the socio-cultural environment [5].

However, like all cross-sectional studies, this research has inherent limitations due to its susceptibility to measurement and reporting biases. Perceptions can vary depending on the context at the time of data collection, which limits the understanding of dynamics following the launch of vaccination campaigns. This underscores the need for a temporal approach to better capture the determinants of vaccine uptake.

Several longitudinal studies have analyzed the evolution of COVID-19 vaccination intent. A study by Latkin et al. (2022) [6] examined the predictors of COVID-19 vaccine uptake in the United States to assess the impact of vaccine hesitancy, social norms, and political affiliation on actual vaccination. Another study by Chambon et al. (2022) [7] analyzed COVID-19 vaccination intent in the Netherlands using longitudinal data collected over six waves between December 2020 and May 2021 to examine the relationships between vaccination intent, attitudes, social norms, and trust over time.

While these two studies provide valuable insights into the evolution of vaccination intent, their analytical approaches have certain limitations. Latkin et al. (2022) [6] use logistic regression, which does not account for temporal variations in vaccination attitudes. Chambon et al. (2022) [7] apply network analysis but do not directly model individual changes over time. By adopting a Generalized Estimating Equations (GEE) modeling approach, this study offers a more robust framework, accounting for intra-individual correlations and enabling a more precise estimation of changes in vaccination intent based on its determinants. This approach is particularly relevant for analyzing the impact of public health policies on the evolution of vaccination attitudes and the adaptation of behaviors in response to vaccination campaigns.

Thus, this research aims to provide a more detailed and dynamic analysis of vaccination intent in West Africa by examining how perceptions, social influences, and health policies interact over time. By leveraging longitudinal data and a methodology suited to repeated measures, it identifies the most significant factors influencing vaccine uptake and offers informed recommendations to strengthen communication and intervention strategies for vaccination. This study will contribute to the advancement of knowledge on vaccine hesitancy and its dynamics while guiding future public health policies aimed at improving vaccination coverage.

## MATERIALS ET METHODS

### Study area

Senegal is a West African country composed of 14 administrative regions. In 2019, the population of Senegal was estimated at 16,209,125 inhabitants, with a relatively balanced distribution (50.2% women and 49.8% men). The ratio of phone numbers per person is 1.1, and the proportion of individuals using a mobile phone at least five times per day increased from 36.4% in 2014 to 73.5% in 2017 [8].

With a population of 11,496,140 inhabitants, of whom 50.9% are women, Benin is also a West African country. In 2021, the mobile phone penetration rate was estimated at 101.8%, corresponding to a mobile subscriber base of 12,731,782 [9].

### Study Design

This is a longitudinal, descriptive, and analytical study. Data collection was conducted in two distinct phases in each country. In Senegal, the first phase took place from December 24, 2020, to January 16, 2021, while the second phase occurred from September 10 to November 26, 2021. In Benin, the first phase, conducted from March 29 to May 14, 2021, and the second phase was carried out from December 15, 2021, to January 14, 2022.

### Study Population

The study targeted adult populations in Senegal and Benin, aged 18 and above, who owned a mobile phone—a necessary requirement due to the telephone-based data collection method.

### Sampling

The sampling was designed to ensure the representativeness of adult populations owning a mobile phone. The sample size was determined using simulations based on the following formula [10, 11]:

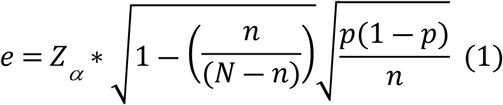

∎ N = Parent population size
∎ n = Sample size
∎ p = Expected proportion in the population
∎ α = Confidence level
∎ *Z_α_* = Value read from the standard normal distribution table

Simulations indicated that a sample of 1,000 individuals provides an approximate precision of 3% when the parent population exceeds 100,000. **Table 2** illustrates the effect of population and sample size on the level of result precision:

**Table 1:**
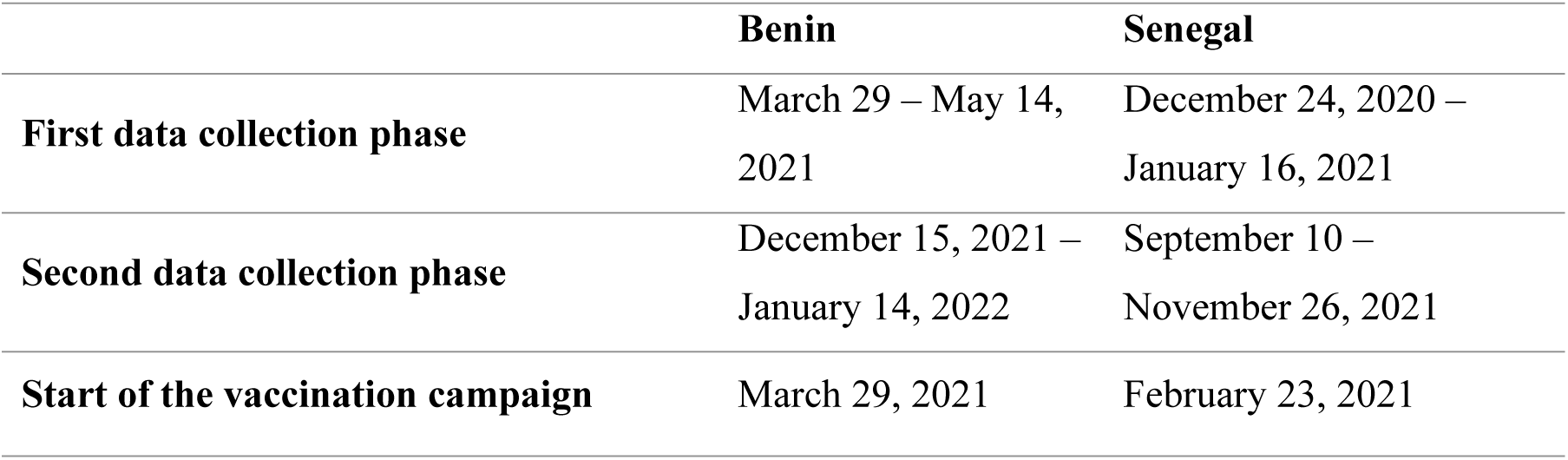
Data Collection Periods and Start of Vaccination Campaigns.

|  | Benin | Senegal |
| --- | --- | --- |
| **First data collection phase** | March 29 – May 14, 2021 | December 24, 2020 – January 16, 2021 |
| **Second data collection phase** | December 15, 2021 – January 14, 2022 | September 10 – November 26, 2021 |
| **Start of the vaccination campaign** | March 29, 2021 | February 23, 2021 |

**Table 2:** Simulations on the Effects of Population and Sample Sizes on Precision.

| N | n | P | $\alpha$ | e |
| --- | --- | --- | --- | --- |
| 10 000 | 1000 | 50% | 5% | 2.9% |
| 50 000 | 1000 | 50% | 5% | 3.1% |
| 100 000 | 1000 | 50% | 5% | 3.1% |
| 200 000 | 1000 | 50% | 5% | 3.1% |
| 500 000 | 1000 | 50% | 5% | 3.1% |
| 1 000 000 | 1000 | 50% | 5% | 3.1% |
| 10 000 000 | 1000 | 50% | 5% | 3.1% |
| 100 000 000 | 1000 | 50% | 5% | 3.1% |

A marginal quota survey was conducted on a target sample of 1,000 individuals [12]. This method is particularly relevant in emergency contexts, such as the COVID-19 pandemic, when sample sizes are below 3,000 [12,13]. A careful selection of quotas can reduce the variance of estimates and narrow confidence intervals. When applied rigorously, quota sampling can provide precision comparable to that of random sampling [13] and may even be superior when the sample size is small [13,14].

During the first phase, the survey questionnaire was administered to a final sample of 813 individuals in Senegal and 865 individuals in Benin. The second phase of data collection followed a longitudinal methodology, targeting only individuals who participated in the first phase and provided their consent to respond again. This process resulted in a final sample of 319 individuals in Senegal and 546 individuals in Benin.

### Theoretical and Conceptual Framework

The study questionnaire was designed based on two well-established theoretical models, deliberately chosen for their direct relevance to the central research question. These two fundamental models are the Theory of Planned Behavior (TPB) and the Health Belief Model (HBM) [5, 15, 16].

The integration of the TPB enables the exploration of complex interactions between individuals’ attitudes, social influence, perceived behavioral control, and their intentions to adopt specific health-related behaviors. This model provides a comprehensive framework for analyzing how personal beliefs, social influences, and perceived control impact behavioral choices.

The use of the HBM enhances our understanding by addressing the perceptual and motivational aspects of health-related decisions. This model emphasizes individuals’ perceptions of the severity of a health issue, their susceptibility to it, the perceived benefits of preventive actions, and the barriers that may hinder these actions. By integrating the HBM, we gain insights into the cognitive processes underlying health-related choices.

By combining these two theoretical models in the questionnaire design, we aim to comprehensively capture the multidimensional aspects of individuals’ decision-making processes regarding health behaviors. This integrated approach allows us to explore not only the motivational factors driving these behaviors but also the cognitive evaluations that influence them. The thoughtful integration of **TPB** and **HBM** into our questionnaire design enhances the depth and precision of our investigation, providing a robust framework for analyzing the factors that shape health-related decisions in the context of our study [5, 15, 16].

## Description of scales and subscales

### Information-seeking behavior on the COVID-19 vaccine

This scale measures proactive information-seeking behavior regarding the COVID-19 vaccine through various means, including regular search behaviors, efforts to better understand the vaccine, and engagement with information received via social media. It is divided into three subscales:

∎ **Regular information search** – Assesses the frequency and consistency of seeking vaccine-related information from various sources (e.g., official websites, media, healthcare professionals).
∎ **Effort to understand the vaccine** – Evaluates the extent to which individuals actively try to comprehend vaccine mechanisms, effectiveness, and potential side effects.
∎ **Engagement with social media information** – Measures how individuals interact with vaccine-related content on social media, including sharing, discussing, or verifying the information received.

Cronbach’s alpha coefficients ranging from **0.68 to 0.71** indicate an acceptable internal consistency for this dimension across both countries and both phases [17, 18] (Table 3). This suggests that the items reliably measure vaccine information-seeking behavior.

**Table 3:** Internal Consistency of COVID-19 vaccination-related dimensions (Cronbach’s Alpha)

| Scale | Subscale | Benin |  | Senegal |  |
| --- | --- | --- | --- | --- | --- |
|  |  | Phase 1 | Phase 2 | Phase 1 | Phase 2 |
| **Information-seeking behavior on the COVID-19 vaccine** | Regular information search | 0.681 | 0.710 | 0.703 | 0.705 |
|  | Effort to understand the vaccine |  |  |  |  |
|  | Engagement with social media information |  |  |  |  |
| **Perception of COVID-19 vaccine safety** | Belief in vaccine safety (reverse-scored) | 0.713 | 0.656 | 0.698 | 0.671 |
|  | Perceived health risk |  |  |  |  |
|  | Concerns about side effects |  |  |  |  |
| **Perceptions of the benefits of COVID-19 vaccination** | Individual protection against the virus | 0.935 | 0.913 | 0.935 | 0.899 |
|  | Reduction of virus transmission |  |  |  |  |
|  | Protection of loved ones |  |  |  |  |
| **Perception of COVID-19 vaccine effectiveness** | Perceived preventive effectiveness against infection | 0.667 | 0.617 | 0.870 | 0.882 |
|  | Risk reduction |  |  |  |  |
| **Social influence on the decision to get vaccinated against COVID-19** | Social pressure from close contacts | 0.864 | 0.772 | 0.871 | 0.803 |
|  | Approval from influential people |  |  |  |  |
|  | Perspective of healthcare professionals |  |  |  |  |
| **Perceived behavioral control** | Ease of access to healthcare professionals | 0.789 | 0.603 | 0.607 | 0.613 |
|  | Trust in healthcare providers |  |  |  |  |
|  | Ease of access to vaccination campaigns |  |  |  |  |
| **Attitudes toward COVID-19 vaccination** | Importance of vaccination | 0.807 | 0.759 | 0.743 | 0.851 |
|  | Usefulness of vaccination |  |  |  |  |
|  | Responsibility toward vaccination |  |  |  |  |
|  | Vaccine Safety |  |  |  |  |
|  | Desirability of vaccination |  |  |  |  |

### Perception of COVID-19 vaccine safety

This scale aims to assess individuals’ perceptions of the safety of the COVID-19 vaccine. It includes three variables measuring different aspects of this perception:

∎ **Belief in vaccine safety (reverse-scored)** – Measures the extent to which individuals trust vaccine developers to ensure its safety. Responses are often reverse scored, meaning higher scores indicate greater confidence in vaccine safety.
∎ **Perceived health risk** – Evaluates whether individuals believe the COVID-19 vaccine poses a threat to their health. Responses indicate the degree to which they perceive the vaccine as potentially dangerous.
∎ **Concerns about side effects** – Assesses individuals’ worries about potential side effects of the COVID-19 vaccine. Responses reflect the level of concern about possible adverse reactions.

With Cronbach’s alpha values ranging from **0.65 to 0.71**, the internal consistency of this dimension is considered acceptable [17, 18] (Table 3). The associated items consistently assess individuals’ perceptions of vaccine safety across both national contexts.

### Perceptions of the benefits of COVID-19 vaccination

This scale assesses how individuals perceive the benefits of COVID-19 vaccination, both personally and at the societal level. It includes three statements, each representing a different aspect of these perceptions:

∎ **Individual protection against the virus** – Evaluates individuals’ perception of the direct personal benefits of vaccination, particularly in terms of protection against infection.
∎ **Reduction of virus transmission** – Measures individuals’ belief in the collective role of vaccination in limiting community transmission and mitigating the impact of the pandemic.
∎ **Protection of loved ones** – Examines the interpersonal perception of vaccination benefits, highlighting individuals’ motivation to protect their close contacts, especially vulnerable individuals.

The highly elevated Cronbach’s alpha coefficients, ranging from **0.90 to 0.94**, demonstrate excellent internal consistency [17, 18] (Table 3). This indicates that the items consistently measure perceived vaccination benefits, reinforcing the validity of this scale in both countries.

### Perception of COVID-19 vaccine effectiveness

This scale assesses how individuals perceive the effectiveness of the COVID-19 vaccine. It includes two statements, each representing a distinct aspect of this perception:

∎ **Perceived preventive effectiveness against infection** – Measures individuals’ belief in the likelihood of being infected with COVID-19 after vaccination.
∎ **Risk reduction** – Evaluates individuals’ perception of the vaccine’s ability to lower the risk of contracting COVID-19. Responses reflect their confidence in the vaccine’s effectiveness in reducing infection risk.

Cronbach’s alpha values ranging from **0.62 to 0.88** indicate internal consistency ranging from acceptable to very good [17, 18] (Table 3). Although slightly lower in Benin, this dimension overall demonstrates satisfactory validity for measuring perceptions of vaccine effectiveness.

### Social influence on the decision to get vaccinated against COVID-19

This scale assesses the perceived influence of social and professional circles on the decision to receive the COVID-19 vaccine. It includes three statements, each representing a distinct aspect of social influence:

∎ **Social pressure from close contacts** – Measures individuals’ perception that, once the COVID-19 vaccine is available, important people in their lives (family, friends) believe they should get vaccinated. Responses reflect the impact of these close contacts’ opinions on their decision.
∎ **Approval from influential people** – Evaluates individuals’ perception that people whose opinions matter to them would support their decision to get vaccinated. It quantifies the influence of this approval on their choice.
∎ **Perspective of healthcare professionals** – Measures individuals’ perception that healthcare professionals would recommend vaccination once the vaccine becomes accessible. It assesses the impact of medical professionals’ opinions on their vaccination decision.

With Cronbach’s alpha values ranging from **0.77 to 0.87**, the internal consistency of this dimension is considered good [17, 18] (Table 3). The items reliably assess the perceived social influence on the decision to get vaccinated in both countries.

### Perceived behavioral control

This scale assesses individuals’ perceptions of their ease of access to COVID-19 vaccination services and their personal autonomy in making vaccination decisions. It includes four statements, each representing a specific aspect of access and decision-making:

∎ **Ease of access to healthcare professionals** – Measures individuals’ perception of their ability to reach healthcare professionals or vaccination centers, influencing their sense of control over the vaccination decision.
∎ **Trust in healthcare providers** – Evaluates the extent to which trust in healthcare providers affects individuals’ perceived control over the vaccination process.
∎ **Ease of access to vaccination campaigns** – Examines individuals’ perception of their ability to participate in organized vaccination campaigns, reducing perceived barriers and increasing their sense of control over the process.
∎ **Personal decision-making** – Assesses individuals’ belief that the decision to receive the vaccine is entirely their own, highlighting their perceived autonomy in the vaccination decision-making process.

After analyzing item-test and item-rest correlations <u>(Table A1)</u>, it was found that the variable “Personal Decision-Making” exhibited weak correlations with the total scale score, suggesting that it did not measure the same construct as the other items. Additionally, an item-test correlation below 0.20 is often considered a threshold below which an item is deemed non-discriminating and may be considered for removal [18]. Consequently, to improve the reliability and validity of the scale, the removal of the “Personal Decision-Making” item was deemed appropriate.

### Attitudes toward COVID-19 vaccination

This scale assesses individuals’ attitudes toward COVID-19 vaccination. It consists of four statements, each reflecting a distinct aspect of these attitudes:

∎ **Importance of vaccination** – Measures individuals’ perception of the significance of getting vaccinated against COVID-19, evaluating the extent to which vaccination is considered a protective measure.
∎ **Usefulness of vaccination** – Assesses individuals’ belief in the effectiveness of vaccination for protecting against COVID-19, reflecting their perception of its benefits.
∎ **Responsibility toward vaccination** – Examines individuals’ perception that getting vaccinated is a responsible act. It measures the sense of duty toward the community.
∎ **Vaccine Safety** – Evaluates individuals’ belief that the COVID-19 vaccine will not pose a health risk, reflecting their perception of vaccine safety.
∎ **Desirability of vaccination** – Measures individuals’ perception of how desirable getting vaccinated against COVID-19 is. It assesses the level of appeal associated with vaccination.

Cronbach’s alpha values ranging from **0.74 to 0.85** indicate good to very good internal consistency [17, 18] (Table 3). This confirms that the items reliably measure individuals’ attitudes toward vaccination in both contexts.

### Data collection and management

A system using the **Random Digit Dialing (RDD)** method [19] was implemented for data collection. The first step involved generating random phone numbers based on the market shares of different mobile operators in each country. Valid numbers were identified by sending bulk SMS messages and analyzing the delivery status. An automated call center then dialed the valid numbers, obtained participants’ consent, and administered the digital questionnaire using ODK software.

After a three-day training on the survey protocol and content, interviews were conducted by enumerators proficient in the main national languages of Benin (Fon, Yoruba, Bariba, Dendi, and Adja) and Senegal (Wolof, Pulaar, Serer, Mandinka, and Jola), in addition to French.

Data Quality Assurance (DQA) was integrated at all stages: before, during, and after data collection. Before data collection, DQA included tool verification, preliminary testing, enumerator selection and training, and ethical approvals to ensure an efficient and standardized process. During data collection, DQA focused on resolving unforeseen issues and adjusting approaches with appropriate guidance. After data collection, DQA involved data alignment, anomaly detection, statistical summaries, pre-post comparisons, and outlier identification using graphical and statistical methods.

To ensure respondent confidentiality, all identifying information, including geographic location, names, and phone numbers, was removed before any data sharing. Only domain-specific identification codes were retained in the electronic data files as part of the anonymization process.

## Data analysis

Data analysis was conducted using GEE models, a suitable approach for longitudinal data due to its ability to account for within-subject correlations. This method is widely used in public health studies to analyze repeated or correlated data while providing robust estimates [20]. It effectively models vaccination intent by considering the correlation between repeated measurements and delivering reliable estimates of the effects of sociodemographic and perceptual variables.

### Dependent variable

Vaccination intent, the dependent variable in this study, was coded dichotomously: 1 indicating a positive intent to get vaccinated and 0 representing a lack of intent. The transformation was carried out by grouping respondents who expressed moderate to strong agreement (“somewhat agree” and “strongly agree”) into the “positive intent” category, while those who expressed opposition or indecision (“somewhat disagree,” “strongly disagree,” and “don’t know”) were classified under “lack of intent.” This dichotomization, a commonly used practice in public health research, facilitates the application of models suited for binary variables [21, 22].

### Independent variables

The independent variables include sociodemographic factors (data collection phase, gender, age, education level) and vaccination-related perceptions (attitudes, perceived benefits, perceived safety, perceived effectiveness, social influence, behavioral control, and information-seeking behavior). This selection aims to provide a comprehensive understanding of the potential determinants of vaccination intent.

### GEE model specification

Given the binary nature of the dependent variable, a binomial distribution with a logit link function was chosen. To determine the most appropriate correlation structure, the Quasi-likelihood Information Criterion (QIC) was used to assess the goodness of fit of different models [23, 24]. A lower QIC value indicates a better balance between model fit and simplicity.

The “Independent” correlation structure appeared to provide a slightly better fit for the data in Benin compared to Senegal. Moreover, the QIC values for different correlation structures were very similar between the two countries, with only minimal differences. This suggests that the choice of correlation structure does not have a significant impact on the overall model fit in these specific cases [23, 24].

**Table 4:**
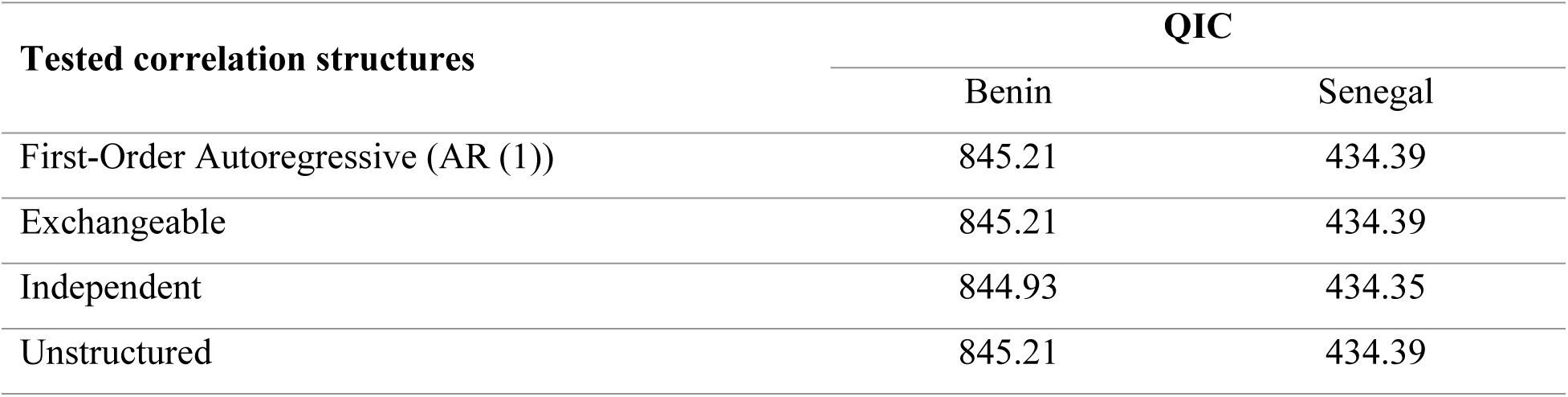
Comparison of Correlation Structures Using the Quasi-likelihood Information Criterion (QIC) for Benin and Senegal.

### Model validation

The GEE model was specified with robust standard errors to ensure reliable statistical inferences, even in the presence of heteroscedasticity or incorrect specifications of the correlation structure [25]. This approach guarantees the validity of statistical tests and confidence intervals without requiring specific tests for heteroscedasticity [25].

A 5-fold cross-validation was also conducted to assess the model’s predictive performance on data not used during estimation [26]. This method randomly divides the data into five subsets and, for each subset, fits the model using the remaining four subgroups while predicting the dependent variable in the unused subgroup. Goodness-of-fit measures, such as the Root Mean Squared Error (RMSE), are then calculated for each iteration.

One approach involves normalizing the RMSE by dividing it by the range of observed values (i.e., the difference between the maximum and minimum values). This normalization produces a value between 0 and 1, where values closer to 0indicate a better model fit.

For Benin, RMSE values range from **0.311 to 0.344**, while for Senegal, they vary between **0.266 and 0.395**. This variation suggests that the predictive performance of the model is acceptable within each country [26].

**Table 5:**
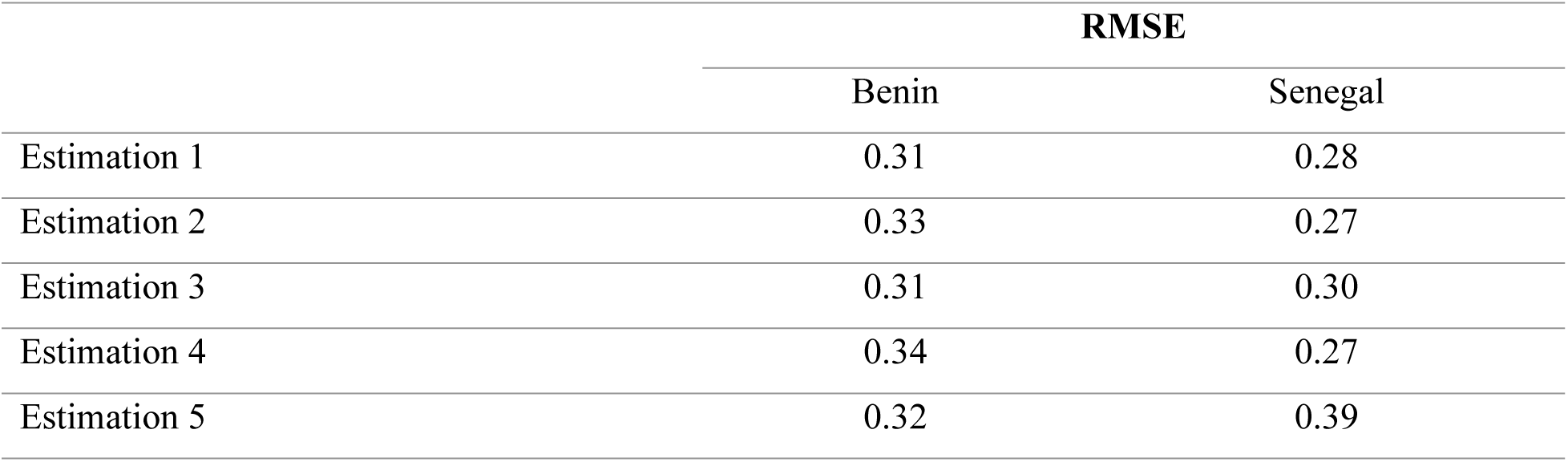
Evaluation of predictive performance using cross-validation.

|  | RMSE |  |
| --- | --- | --- |
|  | Benin | Senegal |
| Estimation 1 | 0.31 | 0.28 |
| Estimation 2 | 0.33 | 0.27 |
| Estimation 3 | 0.31 | 0.30 |
| Estimation 4 | 0.34 | 0.27 |
| Estimation 5 | 0.32 | 0.39 |

### Software

All analyses were conducted using Stata 14, employing the XTGEE command [27] to estimate GEE models with the appropriate options for distribution family, link function, correlation structure, and robust standard error calculation.

### Ethical considerations

The study received approval from the National Ethics Committee for Health Research of Senegal (**SEN/20/23**) and the Local Ethics Committee for Biomedical Research of the University of Parakou in Benin (**0308/CLERB-UP/P/SP/R/SA**). All participants were informed about ethical considerations and their right to withdraw from the study at any time. They all provided their consent to participate.

### Inclusivity in global research

Additional information regarding the ethical, cultural, and scientific considerations specific to inclusivity in global research is included in the Supporting Information.

## RESULTS

### Sample Characteristics

The samples from both countries have a similar gender distribution, with approximately 61% male participants in each case.

Regarding age, Benin is characterized by a predominantly young population, with 56.2% of participants under 25 years old, while in Senegal, this age group represents only 18.5%. Conversely, the 25–59 age group is predominant in Senegal (nearly 70%), compared to less than 38% in Benin.

In terms of education, 63.5% of Beninese participants are educated, a slightly higher percentage than in Senegal (58.3%).

**Table 6:** Sample Characteristics.

|  | Benin |  | Senegal |  |
| --- | --- | --- | --- | --- |
|  | N | % | N | % |
| **Gender** |  |  |  |  |
| Male | 335 | 61.4 | 196 | 61.4 |
| Female | 211 | 38.6 | 123 | 38.6 |
| **Age** |  |  |  |  |
| < 25 years | 307 | 56.2 | 59 | 18.5 |
| 25-59 years | 205 | 37.5 | 223 | 69.9 |
| ≥ 60 years | 34 | 6.2 | 37 | 11.6 |
| **Education** |  |  |  |  |
| Non-educated | 199 | 36.4 | 133 | 41.7 |
| Educated | 347 | 63.5 | 186 | 58.3 |

### Evolution of vaccination intent between the two data collection phases

In Senegal, vaccination intent significantly increased between the two phases, rising from 55.76% to 68.22% (+12.5 percentage points, p = 0.000). This increase was particularly pronounced among women (+19.9 percentage points, p = 0.000) and individuals under 25 years old (+15.6 percentage points, p = 0.030), whereas the increase observed among men (+6.2 percentage points, p = 0.080) and those over 60 years old (+11.1 percentage points, p = 0.770) was not statistically significant.

In Benin, the trend was more mixed, with an overall increase from 64.5% to 69.5% (+5.0 percentage points, p = 0.088), which was not statistically significant. Unlike in Senegal, vaccination intent among women slightly declined (–3.1 percentage points, p = 0.530). However, a significant increase was observed among individuals aged 25–59 years (+11.7 percentage points, p = 0.0112), whereas those under 25 years old exhibited a significant decrease (–11.5 percentage points, p = 0.009), suggesting a decline in vaccine acceptance within this age group.

These findings highlight distinct dynamics between the two countries: in Senegal, there was a uniform and substantial increase, particularly among women and younger individuals, whereas in Benin, the trends were more heterogeneous, with a notable decline in vaccination intent among younger individuals. These differences may be attributed to context-specific factors, such as vaccination awareness campaigns and local perceptions of the vaccine.

**Table 7:** Evolution of vaccination intent between the two data collection phases.

|  | Benin |  |  |  | Senegal |  |  |  |
| --- | --- | --- | --- | --- | --- | --- | --- | --- |
|  | Phase 1 | Phase 2 | Diff. | P-value | Phase 1 | Phase 2 | Diff. | P-value |
| **Gender** |  |  |  |  |  |  |  |  |
| Male | 62.4% | 65.5% | 3.1 | 0.412 | 55.8% | 62.0% | 6.2 | 0.080 |
| Female | 37.6% | 34.5% | -3.1 | 0.530 | 55.7% | 75.6% | 19.9 | 0.000 |
| **Age** |  |  |  |  |  |  |  |  |
| < 25 years | 58.0% | 46.5% | -11.5 | 0.009 | 51.5% | 67.1% | 15.6 | 0.030 |
| 25-59 years | 36.7% | 48.4% | 11.7 | 0.011 | 57.8% | 68.7% | 10.9 | 0.000 |
| ≥ 60 years | 5.2% | 5.0% | -0.2 | 0.972 | 60.0% | 71.1% | 11.1 | 0.770 |
| **Total** | **64.5%** | **69.5%** | **5.0** | **0.088** | **55.8%** | **68.2%** | **12.5** | **0.000** |

### Factors influencing the evolution of vaccination intent in Benin

In Benin, the results indicate that several factors significantly influence the evolution of vaccination intent. The data collection phase emerges as a key determinant, as individuals surveyed during the second phase exhibited a significantly higher likelihood of intending to get vaccinated (OR = 6.955, p < 0.001).

Among latent variables, perceived vaccine safety (OR = 0.781, p < 0.001) is a significant negative factor, suggesting that concerns about vaccine safety reduce vaccination intent. Conversely, positive perceptions of vaccine benefits (OR = 1.191, p = 0.005) and perceived vaccine effectiveness (OR = 1.287, p = 0.005) significantly enhance vaccination intent.

Similarly, the evolution of vaccination intent is significantly shaped by social influence (OR = 1.377, p < 0.001) and increasingly favorable attitudes toward vaccination (OR = 1.266, p < 0.001), highlighting their growing impact on individuals’ decisions to get vaccinated over time.

### Factors influencing the evolution of vaccination intent in Senegal

In Senegal, several factors have significantly shaped the evolution of vaccination intent over time, as revealed by the GEE model. The data collection phase appears as a key determinant, with participants in the second phase displaying a substantially higher likelihood of expressing vaccination intent (OR = 5.021, p < 0.001), highlighting a notable shift in attitudes.

The perception of vaccine benefits (OR = 1.412, p < 0.001) has played an increasingly crucial role, demonstrating that as individuals recognize the personal and collective advantages of vaccination, their intent to get vaccinated strengthens over time. Similarly, social influence (OR = 1.310, p < 0.001) has gained importance, illustrating how the support of close contacts and healthcare professionals has progressively encouraged vaccination decisions.

Conversely, perceived vaccine safety (OR = 0.685, p < 0.001) remains a persistent obstacle, indicating that despite ongoing awareness efforts, concerns about side effects and potential risks continue to hinder the adoption of vaccination. Finally, increasingly positive attitudes toward vaccination (OR = 1.254, p < 0.001) have contributed to reducing hesitancy, suggesting that gradual shifts in perceptions are helping to overcome initial reluctance.

### Differences and similarities between the two countries

Both countries share key common determinants. The data collection phase has a significant effect in both contexts, reflecting a favorable temporal dynamic toward increasing vaccination intent, likely influenced by awareness campaigns or improved information dissemination. Additionally, perceived benefits, social influence, and favorable attitudes are major determining factors in both countries.

However, notable differences emerge. In Benin, perceived vaccine effectiveness (OR = 1.287, p = 0.005) is significant, whereas in Senegal, it has no impact. Furthermore, perceived behavioral control (OR = 1.203, p = 0.003) plays a positive role in Benin but is not significant in Senegal, suggesting differences in perceived access to services or the ability to act. Finally, while perceived vaccine safety is significant in both countries, its effect is slightly more pronounced in Senegal (OR = 0.685 vs. 0.781 in Benin), potentially reflecting context-specific concerns in Senegal.

**Table 8:**
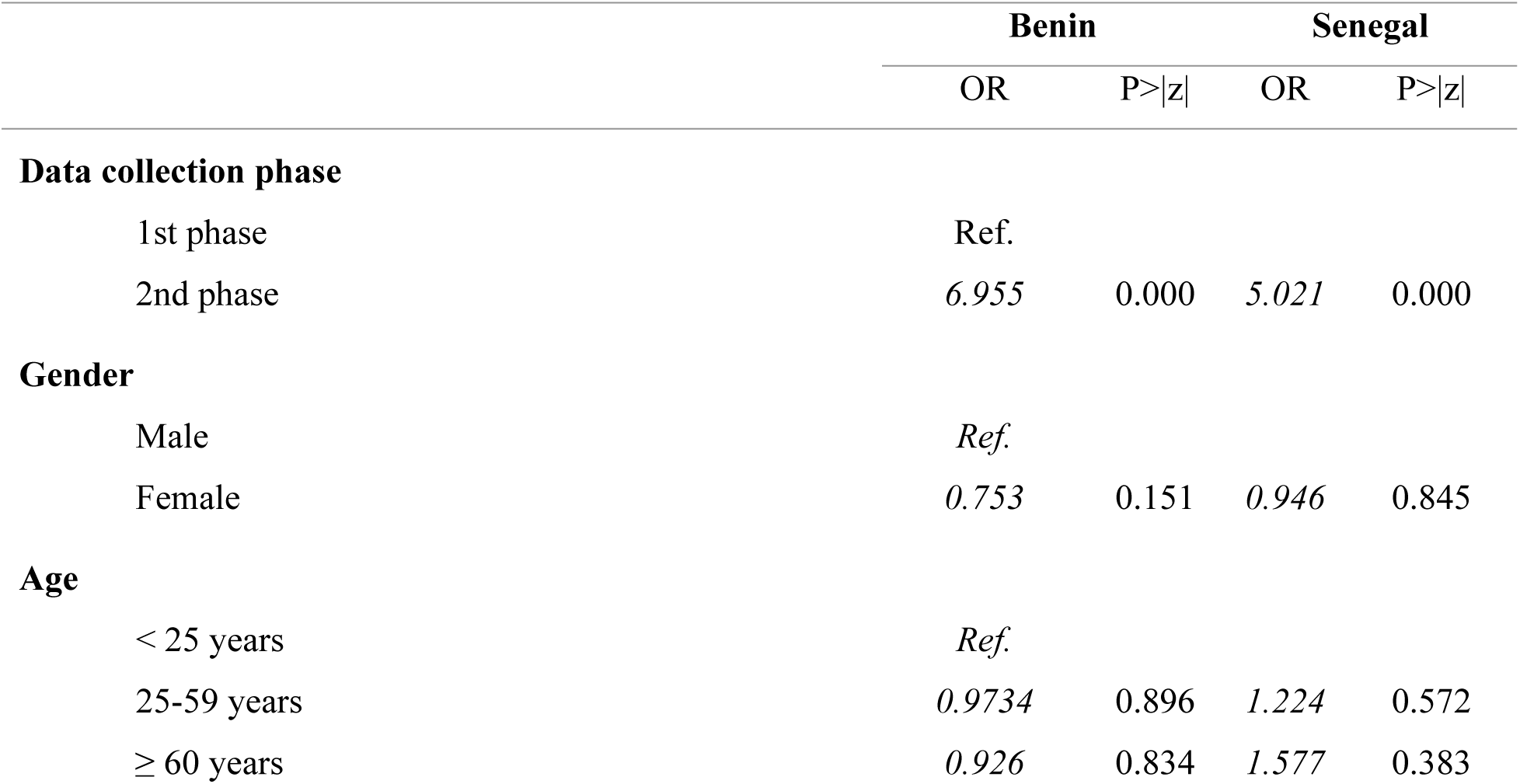
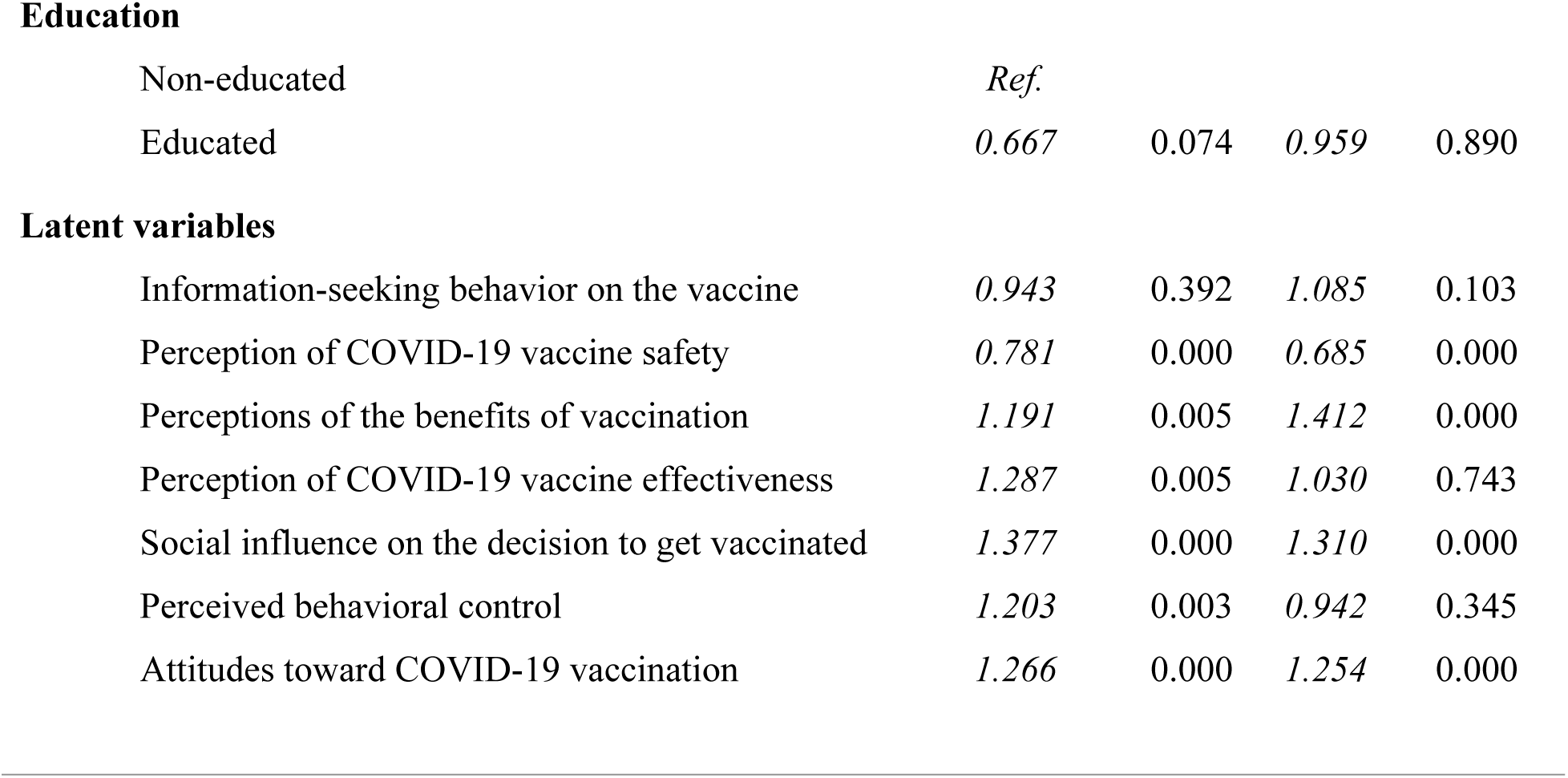
Factors Associated with Vaccination Intent in Benin and Senegal.

|  | Benin |  | Senegal |  |
| --- | --- | --- | --- | --- |
|  | OR | P>\|z\| | OR | P>\|z\| |
| **Data collection phase** |  |  |  |  |
| 1st phase | Ref. |  |  |  |
| 2nd phase | 6.955 | 0.000 | 5.021 | 0.000 |
| **Gender** |  |  |  |  |
| Male | Ref. |  |  |  |
| Female | 0.753 | 0.151 | 0.946 | 0.845 |
| **Age** |  |  |  |  |
| < 25 years | Ref. |  |  |  |
| 25-59 years | 0.9734 | 0.896 | 1.224 | 0.572 |
| ≥ 60 years | 0.926 | 0.834 | 1.577 | 0.383 |
| Education |  |  |  |  |
| Non-educated | *Ref.* |  |  |  |
| Educated | *0.667* | *0.074* | *0.959* | *0.890* |
| Latent variables |  |  |  |  |
| Information-seeking behavior on the vaccine | *0.943* | *0.392* | *1.085* | *0.103* |
| Perception of COVID-19 vaccine safety | *0.781* | *0.000* | *0.685* | *0.000* |
| Perceptions of the benefits of vaccination | *1.191* | *0.005* | *1.412* | *0.000* |
| Perception of COVID-19 vaccine effectiveness | *1.287* | *0.005* | *1.030* | *0.743* |
| Social influence on the decision to get vaccinated | *1.377* | *0.000* | *1.310* | *0.000* |
| Perceived behavioral control | *1.203* | *0.003* | *0.942* | *0.345* |
| Attitudes toward COVID-19 vaccination | *1.266* | *0.000* | *1.254* | *0.000* |

## DISCUSSION

The results reveal that several perceptions significantly influence vaccination intent in both countries, albeit with some notable differences.

In both contexts, **perceived vaccine benefits** play a key role in the evolution of vaccination intent. In both Benin and Senegal, the more individuals recognize the personal and collective advantages of vaccination, the more likely they are to get vaccinated. This finding is consistent with the work of Brewer et al. [28], which demonstrates that emphasizing vaccine benefits can enhance adherence. More broadly, several studies have shown that perceived benefits serve as a central driver in vaccination decision-making. For instance, Rosenstock et al. [29], within the framework of the HBM, emphasize that the more an individual perceives the vaccine as an effective means of protection against a serious disease, the more likely they are to adopt a favorable vaccination behavior. Similarly, Xiao et al. [30], through a meta-analysis on the determinants of COVID-19 vaccine hesitancy, found that highlighting benefits, particularly in terms of individual and collective protection, plays a key role in vaccine acceptance. These observations indicate that effective awareness campaigns should focus on communicating the tangible benefits of vaccination, both at the individual and community levels, to strengthen trust and promote adherence.

**Perceived vaccine safety** constitutes a major barrier in both countries, with a slightly stronger effect in Senegal than in Benin. An increase in concerns regarding side effects and perceived risks hinders vaccination intent, as also reported by Loomba et al. [31]. Similarly, our previous findings [5] confirmed the significant impact of safety perceptions on vaccination intent. The observed difference between Benin and Senegal may be explained by variations in the dissemination and perception of vaccine safety information or by local experiences related to vaccine administration. Indeed, a study revealed that COVID-19 vaccines are perceived as less safe and effective than other vaccines, primarily due to concerns regarding their rapid development and potential side effects [32]. In Senegal, a study identified that the lack of pharmacovigilance data and distrust toward official sources of information contribute to vaccine hesitancy [33]. These findings suggest that vaccine safety perceptions are shaped by contextual factors, such as trust in health authorities, access to reliable information, and past experiences with vaccination programs.

**Social influence** emerges as a key factor in both countries, although its effect is slightly more pronounced in Benin than in Senegal. Greater approval from close contacts, healthcare professionals, and community leaders appears to motivate individuals to get vaccinated, in line with the findings of Omer et al. [34]. Anthropological studies in the region further support this observation. A survey conducted in Benin revealed that vaccine acceptance is strongly influenced by social engagement, particularly through support from healthcare professionals and traditional healers [35]. The findings indicate that vaccine hesitancy is often linked to local beliefs and distrust in government management, suggesting that the involvement of community actors is essential to improving vaccine uptake [35]. In Senegal, a study [5] demonstrated that social interactions and positive community norms promote vaccination adoption. These results suggest that engaging community networks, including local leaders and healthcare professionals, constitutes an effective strategy to increase vaccination intent. By mobilizing these influential actors, it is possible to strengthen trust in vaccines, foster positive attitudes toward vaccination, and ultimately contribute to higher vaccine coverage in the region.

Overall **favorable attitudes toward vaccination** significantly influence the evolution of vaccination intent in both countries. These findings align with numerous studies highlighting the role of positive perceptions in vaccine decision-making. Greyling and Rossouw (2022) [36] demonstrated, through a sentiment analysis of social media discussions across ten countries, that positive attitudes toward vaccination tended to decline over time. Additionally, a systematic review of COVID-19 vaccine attitudes in Africa found that acceptance was driven by trust in government measures against COVID-19 and by personal experiences, such as having close contacts diagnosed with or deceased from the disease [37]. Another study revealed that attitudes and beliefs about vaccines in general, and the COVID-19 vaccine in particular, influence vaccination acceptance [38]. These findings suggest that to strengthen vaccination intent, it is essential to promote the development of positive attitudes toward vaccination. This requires proactive and transparent communication, particularly through social media, to maintain and reinforce favorable attitudes by emphasizing vaccine safety and benefits.

A notable difference lies in the perception of vaccine effectiveness, which is significant in Benin (OR = 1.287, p = 0.005) but not in Senegal. Studies such as those by Brewer et al. [28] have emphasized that perceived vaccine effectiveness is a key factor in the evolution of vaccination intent, but its impact can vary depending on social and cultural contexts.

Recent research in the region provides further insights into these differences. In Benin, a study conducted in April 2021suggests that perceived vaccine effectiveness is influenced by trust in its safety and by the quality of information received [39]. In Senegal, a study conducted in late 2020 found that one of the main reasons for vaccine hesitancy was doubt about its effectiveness, driven by concerns that the vaccine might pose health risks [40]. These findings suggest that, to enhance vaccine acceptance, it is crucial to develop context-specific communication strategies, strengthening transparency and the reliability of information on vaccine effectiveness while considering the sociocultural specificities of each country.

Furthermore, perceived behavioral control (OR = 1.203, p = 0.003), an indicator of the perceived ability to access vaccination services, is significant only in Benin. This divergence may be linked to contextual differences in perceived vaccine access and available resources, as suggested by the work of Galanis et al. [41], who demonstrated that logistical barriers and limited access to services influence vaccination behaviors. This perspective is also supported by Lin et al. [42], who emphasize the importance of improving accessibility to overcome perceived barriers and increase vaccine uptake. These findings indicate that to enhance vaccination adherence in Benin, it is crucial to strengthen perceived accessibility by reducing logistical barriers and improving vaccine distribution.

## LIMITS

The samples were nationally representative only and did not allow for disaggregation by place of residence or region. Moreover, only individuals with a mobile phone were surveyed, thereby excluding the most marginalized populations.

Additionally, it is generally preferable to include at least three items per scale, as was the case for all measured dimensions except for the perception of COVID-19 vaccine effectiveness.

## CONCLUSION

The findings of this study highlight both common and specific determinants of vaccination intent in Benin and Senegal. In both countries, perceived vaccine benefits, social influence, and positive attitudes emerge as key drivers of vaccine uptake. However, contextual differences are noteworthy: in Benin, perceived vaccine effectiveness and behavioral control play a significant role, whereas in Senegal, these factors are not determinants. Additionally, while perceived vaccine safety acts as a barrier in both contexts, its impact is more pronounced in Senegal. These results underscore the need to tailor communication and intervention strategies to the specific realities of each country to maximize vaccine acceptance.

These conclusions align with the work of Dubé et al. (2013) [43], who emphasized the importance of public trust in vaccines and health authorities to ensure the success of vaccination programs. They recommend communication strategies adapted to sociocultural contexts to address population concerns and strengthen vaccine uptake. Furthermore, Dubé and MacDonald (2022) [44] highlighted that understanding and addressing the concerns of hesitant individuals is essential to increasing vaccine acceptance and adoption.

Thus, to improve vaccine uptake, it is crucial to implement clear and context-specific communication strategies, enhance transparency and trust in health systems, and actively engage community leaders and healthcare professionals. These approaches will enable an effective response to public concerns and maximize adherence to vaccination programs.

## CONFLICT OF INTEREST

- The authors declare that they have no competing financial interests.
- There are no competing interests related to this work.
- There are no known conflicts of interest associated with this publication.

All authors attest they meet the ICMJE criteria for authorship.

## FINANCING

This research is part of the African Response to the COVID-19 Epidemic Support Program (ARIACOV), funded by the French Development Agency (AFD). The funding organizations played no role in the study design, data collection and analysis, decision to publish, or manuscript preparation.

## Data Availability

GAYE, Ibrahima (2025). Stata database. figshare. Dataset. https://doi.org/10.6084/m9.figshare.28717388.v1 GAYE, Ibrahima (2025). Stata analysis code. figshare. Dataset. https://doi.org/10.6084/m9.figshare.28717391.v1

https://doi.org/10.6084/m9.figshare.28612238.v1

## APPENDICES

**Table A1:** Evaluation of the Impact of Items on the Internal Consistency of the Dimension.

|  | Item-test correlation |  |  |  | Item-rest correlation |  |  |  |
| --- | --- | --- | --- | --- | --- | --- | --- | --- |
|  | Benin |  | Senegal |  | Benin |  | Senegal |  |
|  | Phase 1 | Phase 2 | Phase 1 | Phase 2 | Phase 1 | Phase 2 | Phase 1 | Phase 2 |
| Ease of access to healthcare professionals | 0.82 | 0.66 | 0.60 | 0.72 | 0.65 | 0.24 | 0.30 | 0.40 |
| Trust in healthcare providers | 0.81 | 0.68 | 0.72 | 0.62 | 0.58 | 0.34 | 0.40 | 0.33 |
| Ease of access to vaccination campaigns | 0.80 | 0.61 | 0.80 | 0.80 | 0.61 | 0.31 | 0.56 | 0.54 |
| Personal decision-making | 0.50 | 0.48 | 0.53 | 0.55 | 0.25 | 0.11 | 0.24 | 0.28 |

